# Plugging the holes in the Swiss Cheese by learning from those who fall through the cracks of our medical system’s multiple lines of defense in Hong Kong: Data-based analytic protocol for identifying the tertiary prevention needs of a population with machine learning pipeline

**DOI:** 10.1101/2022.06.20.22276619

**Authors:** Eman Leung, Albert Lee, Jingjing Guan, Chun Cheung Ching, Olivia Lam, Bonnie Wong, Chun Bon Law, Hector Tsang

**Author notes:** Corresponding author Correspondence: Jingjing Guan. Address: 4/F, The Jockey Club School of Public Health and Primary Care, The Chinese University of Hong Kong, Prince of Wales Hospital, Sha Tin, Hong Kong Special Administrative Region of the People’s Republic of China.

## Abstract

**Introduction:** The objective of tertiary prevention is to reduce re-hospitalization, as re-hospitalization puts patients at unnecessary risk, delays those in the population requiring timely care, and incurs financial burdens on healthcare systems. Nevertheless, it is challenging to stratify the needs of tertiary prevention in a population. Hence, we advance an analytic protocol to identify from an inpatient population the clinical and service-utilization profiles of those re-hospitalized within 28 days.

**Methods and analysis:** The protocol is based on implementing unsupervised and supervised machine learning (ML) in tandem with an inpatient population’s electronic health records. The unsupervised ML will cluster the population into segments of maximized within-segment similarity and between-segment dissimilarity, across the dimensions of clinical diagnoses, acuity, complexity, chronicity, and multimorbidity.

Within each clinically similar segment identified, a 28-day re-hospitalization outcome-supervised decision tree will classify the segment into a series of binarily-split subgroups with profile and service utilization-related features. The order of selected features reflects relative importance to the outcome. Two subgroups originated from a selected feature are statistically different in re-hospitalization outcomes. So, the subgroups lacking selected services while realizing the highest re-hospitalization rates will potentially benefit most from tertiary prevention of the selected services. Thus, they are fit for further assessment and corresponding interventions.

**Ethics and dissemination:** The Survey and Behaviour Research Ethics Committee of the Chinese University of Hong Kong, Hospital Authority Data Collaboration Lab, and a local ethics committee have approved this protocol and its ongoing and forthcoming validations in territory-wide HK through centralized data access and in local clinical management systems, respectively. In addition to disseminating through publications, presentations, and other communications, the protocol is also implementable in different systems as part of the decision support mechanism to inform the venue-based sampling of patients with a high risk of re-hospitalization.

**Article Summary:** *Strengths and Limitations of this study:* Strength #1
This will be a data-based machine-learning analytic methodology to align population-based cohort study with the conceptual framework of the Swiss Cheese Model for patient safety. The Swiss Cheese Model is a conceptual framework that elevates us from the paradigm of linear causality to conceptualizing safety events as having multiple lines of defense simultaneously broken through. However, the predominant linear model research is incompatible with identifying a medical system’s multiple lines of defense and their potential breakpoints, due to much less prioritizing impacts and mapping interactions on outcomes. Strength #2
This protocol no long assumes independence among clinical profiles and service utilization-related factors. As a result, the absence and ensembles of post-acute services of the entire population can be handled appropriately. Strength #3
Subgroups of statistically significant differences in re-hospitalization outcomes will be identified and articulable as a portfolio of clinical profiles and selected services. Limitations
Heterogeneity may still exist after the clustering and classification based on clinical homogeneity. Like all studies based on clinical databases, the heterogeneity can be attributable to exogenous factors absent in clinical information systems, e.g., psychosocial factors, and the narrowly defined tertiary prevention needs of the supervisory outcome.

## INTRODUCTION

### Background and rationale

A block of Swiss cheese can be seen through only when all its eyes are aligned. Similarly, Healthcare systems’ safety incidents occur when the systems’ multiple lines of defense are all broken through (Reason, 2000). Notably, the Swiss cheese model has been adopted as a paradigm for investigating the root causes of patient safety incidents across the care continuum from pre-operative consultation to rehabilitation (Zecevic, Li, Ngo, Halligan, & Kothari, 2017; Sankaran, Andrews, Chicas, Wachter, & Berger, 2020). Recently, the Swiss cheese model has also been applied to conceptualize the outbreak of COVID-19 as a reflection that different lines of defense in primary care prevention have been broken through (Noh et al., 2020). Consequently, the Swiss Cheese model has broadened the search for root causes of patient safety events from clinical factors alone to factors associated with resources, culture, health system policies (Waring, Allen, Braithwaite, & Sandall, 2016), and even the cognitive process of clinical decision making (Seshia et al., 2018).

One patient safety outcome receiving the most attention is a patient’s 28-day re-hospitalization (e.g., Hansen, Williams, & Singer, 2010). Conventionally, 28-day re-hospitalizations have been attributed to either the poor care quality that patients receive at acute care hospitals (Zhou et al., 2016) or patients’ clinical characteristics (Mahmoudi et al., 2020).

However, there is a growing recognition that what precipitates 28-day re-hospitalization is multifactorial, including clinical factors as well as medical services utilization, and is, therefore, best understood in terms of the multiple broken lines of defense of the Swiss Cheese model (Seshia et al., 2018). Nevertheless, there lacks any systematic examination of the effects of acute-post-acute and post-acute-post-acute service interactions and the effects of clinical profile-service interactions on re-hospitalizations. Previous studies did examine the relationships between the quality of acute care and the effect of post-acute care on patient outcomes (Kripalani et al., 2014; Chen et al., 2010), between post-acute care and patient safety (Wang, Diamantidis, Wylie, & Greer, 2017; Lafortune, Béland, Bergman, & Ankri, 2009), and between post-acute care and re-hospitalization (Grabowski & Maddox, 2020). It has even been demonstrated with a two-stage analysis that the effect of a specific post-acute service on 28-day re-hospitalization is conditional on patients’ levels of clinical risk (Howard et al., 2021). The literature has yet to model the complex clinical reality wherein patients are discharged with multiple post-acute services depending on acute care quality, patients’ clinical needs, and availability, all of which affect re-hospitalization outcomes.

The traditional analytic approach limits the extent to which current tertiary prevention/re-hospitalization research is congruent with the Swiss Cheese model. Specifically, the prevalent statistical modeling technique in the literature is incompatible with the need to identify healthcare systems’ critical lines of defense for different clinical profiles from an intricate web of relationships among a vast volume of data and features. The dependence among features would invalidate the assumptions of many statistical approaches as well. We noticed that machine learning (ML) with proper design could address these issues.

### Objectives

Reducing re-hospitalization is an objective of tertiary prevention. We will advance an ML-based analytic protocol for identifying the tertiary prevention targets of a regional population from the clinical and service utilization profiles of its residents who were re-hospitalized within 28 days of previous discharges. The specific aim is to design and validate an ML methodology to align with Reason’s (2000) conceptual framework and identify the “multiple lines of defense” in the medical system against re-hospitalization, and profile patients who fall through the cracks of the broken lines of defense.

The writing of this protocol was concurrently driven by our Strategic Public Policy Research (SPPR) research project, which is a peer-reviewed project funding scheme by the HK government’s Policy and Innovation Co-ordinating Office. The current protocol is approved by the SPPR as part of the research design and deliverables. SPPR also funded the consequential implementation of the current protocol in a local healthcare system, as the base to create a sample frame for profiling discharging patient cohort who “fall through the crack” of Swiss Cheese. By the time of writing this protocol, the ethics committee of the intended local healthcare system had just approved (1) the implementation of the current protocol in their regional clinical management system (CMS), (2) to identify patients for in-depth assessment, and (3) to tailor local clinical resources to follow those patients who will likely fall through the cracks. The local healthcare system expects to develop a routine mechanism for post-acute care assignment based on the tagging of high tertiary prevention risk individuals, following the current protocol. It is essential to validate and implement the reporting methodology as standard operating procedures.

The current protocol is under ongoing validation exercises planned in the entire HK at the time of writing. One ongoing validation utilizes the more than 20 years of electronic health records (EHRs) of HK’s territory-wide public health systems. The data access is granted by our collaboration project with the Hospital Authority Data Collaboration Laboratory (HADCL) of the Hospital Authority (HA) of HK, which hosts the clinical information systems of the entire HK’s public medical services. The ongoing validation differs from the implementation conducted in a local healthcare system. As the information infrastructure differs, access rights to data differ from HADCL to the local CMS. Most importantly, the local healthcare system has the authority to allow the implementation and validation of the current protocol in their local CMS, as well as trigger appropriate follow-up care for patients in need.

As a result, it is necessary to establish a consistent set of procedures for the analytics. The protocol will facilitate an informative comparison of the results of the separate validation exercises between the entire HK via HADCL’s centralized data access and the local healthcare system that operates its own CMS. During the ongoing process of validations being conducted in HADCL and the local healthcare system’s CMS, we are requested to showcase the protocol with a small subset of data to explain the protocol, especially the ML pipeline and analysis to non-ML experts. The showcase turned out to be an effective illustration to the general audience. Hence, we include its data-based illustrative example to explain the ML pipeline in this study protocol to facilitate interpretation.

## METHODS AND ANALYSIS

### Study design

As informed by the Swiss Cheese model, the current protocol takes a data-driven approach to identify the critical lines of defense against 28-day re-hospitalization among acute and post-acute services in a medical system. Further, just as a block of Swiss Cheese can be seen through when all its eyes are aligned, re-hospitalizations occur when specific clinical profiles are confronted with gaps in critical post-acute services. Specifically, our unique ML pipeline will identify the system’s key lines of defense and their corresponding gaps through which patients of different clinical profiles may fall. Consequently, our analytic protocol ensures that our research in re-hospitalization within the context of tertiary prevention is congruent with the Swiss Cheese model.

Only an ML pipeline can address the following issues. On the one hand, we need to find clinically homogeneous patients to compare different ensembles of post-acute services. Clustering a population’s EHRs is challenging even with the commonly-used clustering algorithm, given the large volume of patients in the population and features to be examined. In addition, most algorithms cannot cluster with categorical features, not to mention mixed feature types (continuous and categorical), which are common in healthcare research. On the other hand, EHRs’ vast volume of features is not entirely independent from one another. Some clinical features are more likely to co-occur among patients with certain profiles than others. Some features representing specific levels or types of services are also more likely to be associated with specific clinical features. The lack of independence among some features aligns with the clinical reality that service assignment decisions are primarily based on patients’ clinical profiles under the constraints of service availability. In any event, the potential endogeneity resulting from such non-independence can invalidate traditional modeling approaches.

Moreover, a patient can receive none, one, or multiple post-acute services as recorded. The vast volume of patients and their EHRs engendered a large number of “logically missing” data, in addition to random missing ones. Thus, not only can non-random missing data challenge the assumptions of the traditional modeling, but the sparsity resulting from the sheer volume of missing data can also be challenging. The vast volume of features in EHRs also precipitates another kind of sparsity associated with a large number of zeros or values that lack significant impact. Notably, sparsity resulting from non-missing values that are either zero or lacking impact compromises the validity of both ML and traditional statistical models. For example, sparsity may result in a regression-based decision tree with great width but lacks sufficient depth that makes the result interpretable. Features sparsity can result in an overfitted model, whereby noises in the training data contribute to the model fit.

Our ML pipeline will address these issues systematically by grouping clinically similar patients from a population into the same segments and then comparing within each clinically homogeneous segment its patients’ 28-day re-hospitalization outcomes associated with having received different levels of acute care or types of post-acute services. To this end, the objective of the current protocol is concretized to identify the epidemiological and clinical profiles of the subgroup from each clinically homogeneous segment that lacks any service of importance. Please refer to Figure 1 for a schematic presentation of the methodology feature in the current protocol.

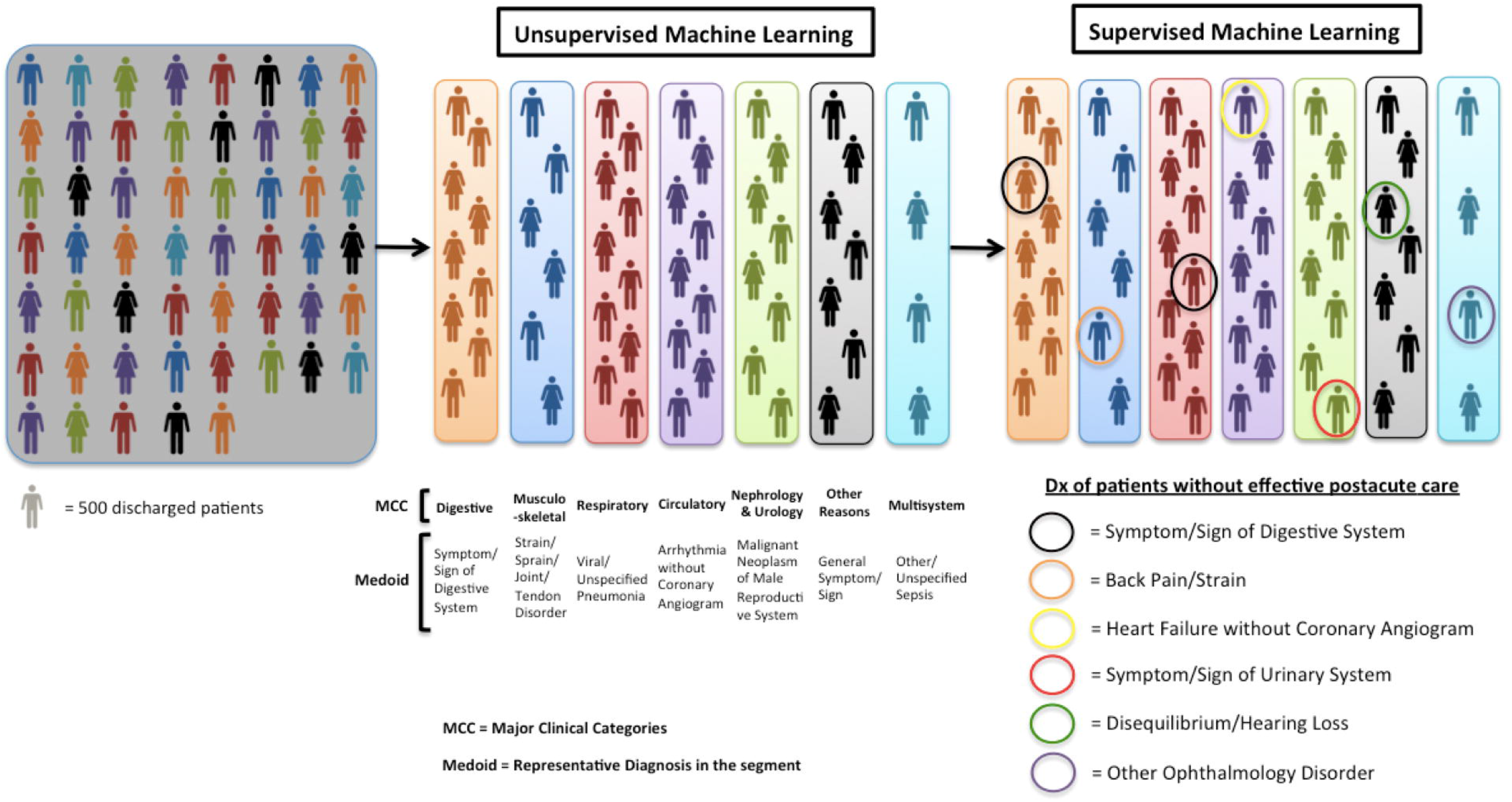

### The machine learning pipeline I: Clustering

The patient population will be first clustered into clinically homogeneous segments to maximize intra-segment similarity and inter-segment dissimilarity according to the following factors with unsupervised (bottom-up) ML: Clinical diagnoses, acuity, severity, chronicity, and resource-intensifying comorbidity (CIHI, 2015). We will use the unsupervised ML algorithm CLARA (Clustering Large Applications; Kaufman & Rousseeuw, 1990) to overcome the healthcare research-relevant technical challenges mentioned above. Furthermore, we will apply an objective metric to select the optimal number of segments that maximize the within-segment homogeneity and between-segment heterogeneity of the studied population (The Calinski-Harabasz Index; Caliński & Harabasz, 1974).

### The machine learning pipeline II: Classification

After grouping the population into segments with clinical profiles, a classification tree algorithm will further include service-related features to partition each clinically homogeneous segment into subgroups. The classification will be supervised by the binary 28-day re-hospitalization outcomes. We will use a non-regression-based decision tree to perform unbiased recursive partitioning and surrogate splitting (URPSS; Hothorn, Hornik, & Zeileis, 2004). On the one hand, the surrogate splitting enables the unbiased comparison between two sides of the splits despite missing data. Each segment will be partitioned with features selected sequentially in an order that reflects the magnitude of their marginal contributions to re-hospitalization outcomes with every feature that remains with variability in the pool. The greater the magnitude of the selected service’s marginal contribution to the 28-day re-hospitalization outcome, the earlier it will be selected by URPSS. Each selected feature (a parental node) will split the remaining records in the segment into two subsets (offspring nodes). On the other hand, the split-stopping criteria of URPSS optimize the tree depth by stopping when the conditional inference of any remaining features in the pool makes no additional marginal contribution to the outcome. It is the URPSS’s property that two offspring nodes split from the same parental node are of statistically significant difference in their outcomes. In other words, the last subsets remaining from the sequential partitioning, i.e., the offspring nodes without turning into parental nodes, can be inferred as no longer affected by any further services that cause significantly different re-hospitalization outcomes.

### The machine learning pipeline III: Analysis and inference

The relative importance of each level/type of service to patients’ 28-day re-hospitalization outcomes can be deduced from the different services that clinically similar patients receive. We hypothesize that those who lack any service of importance to other clinically similar patients of the same segment will exhibit the highest re-hospitalization rate as a subgroup. The patients who fall through the crack as conceptualized in the Swiss Cheese model will be the subgroups of patients that have not utilized any service selected by the classification tree while resulting in the highest 28-day re-hospitalization rate. Till this step, the current protocol allows the data and ML analytics to guide the profiling of the subgroup of patients who did not receive any of the services associated with a significantly lower re-hospitalization rate among clinically similar patients. Detailed analysis and inference approaches will be included in the illustrative example. Please refer to Table 1 for the clinical information in the clustering and classification.

**Table 1.**
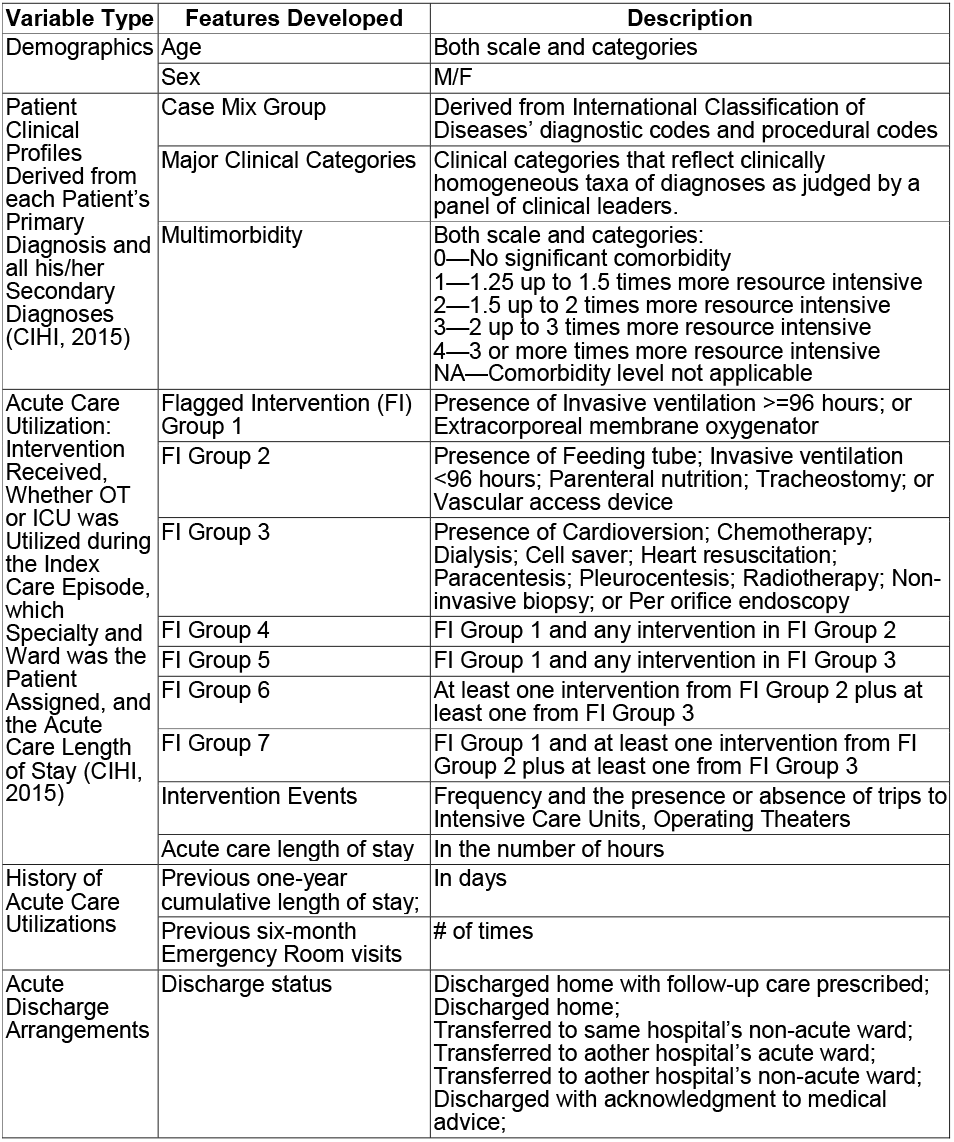
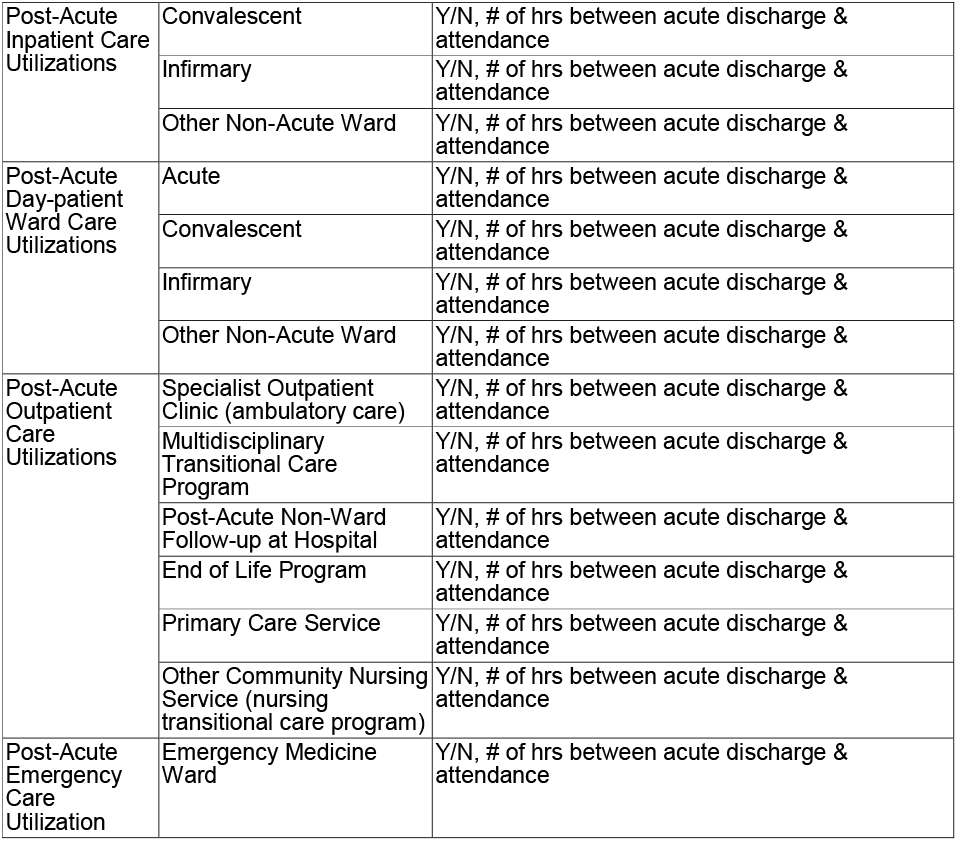
The list of clinical and utilization-related features based on which the protocol was built and the illustrative data analysis was performe

### Study setting, interventions, and eligibility criteria

The proposed protocol can study any patient populations that utilize the acute inpatient services of HK’s public healthcare system. Most acute inpatients in HK were admitted to wards through accidents and emergency departments. The analytic protocol will be tailored to acute inpatients of all age groups and to those who are community-dwelling or residing in long-term care facilities, respectively.

A wide range of post-acute follow-up services has been available for years, including those provided by acute hospitals, convalescent hospitals, specialist outpatient clinics, community nursing, and other doctor services. The current analytic protocol will consider all types of medical services received by eligible patients during their index hospitalization’s post-discharge period and before the subsequent index hospitalization, i.e., the interventions, to ensure the ML-based analytics’ adherence to the set of studied interventions. The assignment of post-acute services is upon a patient’s discharge from acute wards. In order to observe the complete patient journey and ensure the completeness of care episodes, patients who were ever discharged from acute wards with unknown discharge status or against medical advice shall be excluded from the analyzable population.

### Illustrative example

In order to prove the concept, we followed the protocol and conducted an illustrative instance of ML pipeline on community-dwelling inpatients restricted to two senior age groups, i.e., aged 50-64 and 65+ years old, residing in an administrative district in HK and admitted to acute inpatient services between January 1^st^, 2017 to December 31^st^, 2017. Their EHRs were made available by the HADCL.

The CLARA, i.e., the clustering step, partitioned the inpatient EHRs for both 50–64 (n=11,555) and 65+ (n=26,827) respectively into six and seven clinically homogeneous segments. As shown in Table 2, we will report the title based on the Major Clinical Category (MCC; CIHI, 2015) of each segment, as well as its data-based medoid diagnosis. MCC is a standardized categorization of clinical inpatient diagnosis groups. Medoid diagnosis is computationally determined by the data and CLARA. Medoid diagnosis will be selected from the medoid of vectors of input features, whose sum of dissimilarities to all the vectors of input features in the segment is minimal. In turn, medoid diagnosis is a representative diagnosis of that segment. The most representative diagnosis for 50-64 and 65+ samples was Viral/Unspecified Pneumonia. The dominant MCC, medoid diagnoses, segment sizes, and re-hospitalization rates of the optimal composition of patient segments identified by the CLARA clustering will be summarized as Table 2.

**Table 2.** Illustrative table of CLARA clustering outcome of segments from a study population with characteristics of Major Clinical Category, medoid diagnosis, prevalence, and 28-day re-hospitalization rate for two study populations (50-64 and 65+).

|  | Segment | Age = 50 - 64 | Segment | Age = 65+ |
| --- | --- | --- | --- | --- |
| 1 | **Respiratory System**<br>(Medoid diagnosis: Viral/Unspecified Pneumonia) | n = 1235<br>rate = 24.78% | **Respiratory System**<br>(Medoid diagnosis: Viral/Unspecified Pneumonia) | n = 4864<br>rate = 23.56% |
| 2 | **Nephrology & Urology**<br>(Medoid diagnosis: Urinary Obstruction without Percutaneous Drainage) | n = 2761<br>rate = 24.74% | **Nephrology & Urology**<br>(Medoid diagnosis: Malignant Neoplasm of Male Reproductive System) | n = 4137<br>rate = 24.08% |
| 3 | **Digestive System**<br>(Medoid diagnosis: Other Gastrointestinal Disorder) | n = 2207<br>rate<sup>†</sup> = 19.66% | **Digestive System**<br>(Medoid diagnosis: Symptom/ Sign of Digestive System) | n = 4566<br>rate = 20.28% |
| 4 | **Circulatory System**<br>(Medoid diagnosis: Unstable Angina/Atherosclerotic Heart Disease without Coronary Angiogram) | n = 1505<br>rate = 17.67% | **Circulatory System**<br>(Medoid diagnosis: Arrhythmia without Coronary Angiogram) | n = 4317<br>rate = 21.66% |
| 5 | **Musculoskeletal Systems & Connective Tissue**<br>(Medoid diagnosis: Inflammatory and Reactive Arthropathy) | n = 1879<br>rate = 12.03% | **Musculoskeletal Systems & Connective Tissue**<br>(Medoid diagnosis: Strain/Sprain/Joint/Tendon Disorder) | n = 3147<br>rate = 13.50% |
| 6 | **Other Reasons for Hospitalization**<br>(Medoid diagnosis: General Symptom/ Sign) | n = 1968 | **Other Reasons for Hospitalization**<br>(Medoid diagnosis: General Symptom/ Sign) | n = 3572 |
|  |  | rate = 11.89% |  | rate = 13.38% |
| 7 | - | - | **Multisystem-Unspecific Site Infection and Blood Disorder**<br>(Medoid diagnosis: Other/ Unspecified Sepsis) | n = 2224 |
|  |  | - |  | rate = 17.54% |
|  | **Overall** | n = 11555 | **Overall** | n = 26827 |
|  |  | rate = 18.60% |  | rate = 19.74% |

<sup>†</sup> rate = re-hospitalization rate of the respective segment

After the clustering, the URPSS decision tree classified each CLARA-based segment’s records according to 28-day re-hospitalization outcomes. Using the respiratory segments for the 50-64 and 65+ study population, Figures 2a and 2b are the schematics of what a within-segment URPSS ML analysis will show. Each schematic will have the overall segment at the top of the diagram. Then, according to the features selected in an order that reflects each feature’s marginal contribution to 28-day re-hospitalization, the schematic will split into a hierarchical structure with corresponding features as nodes. Each node’s sample size and 28-day re-hospitalization outcomes will be listed. The protocol will focus on showing the selected service features, by subtracting a decision tree’s diagram to anchor on the service features.

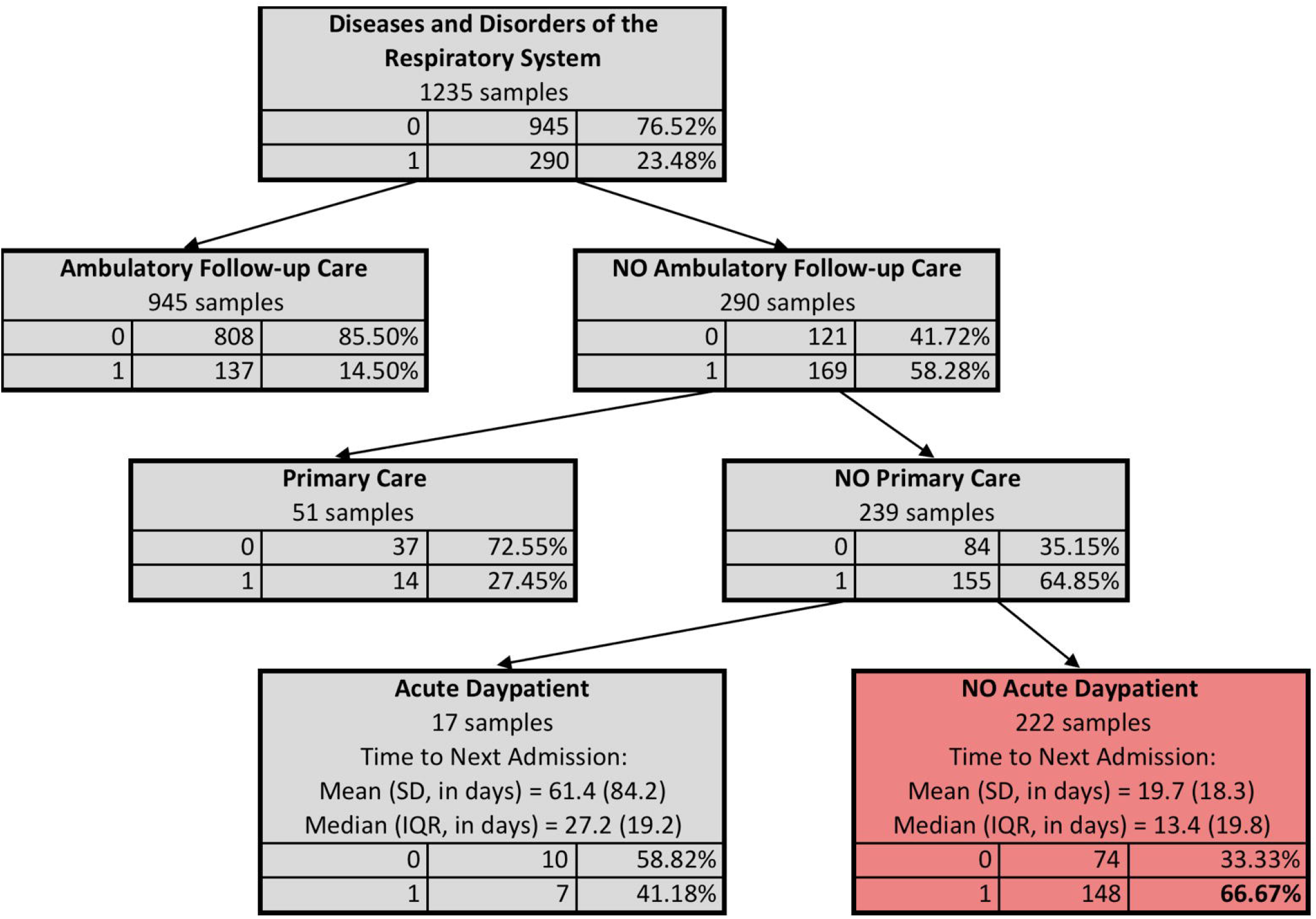

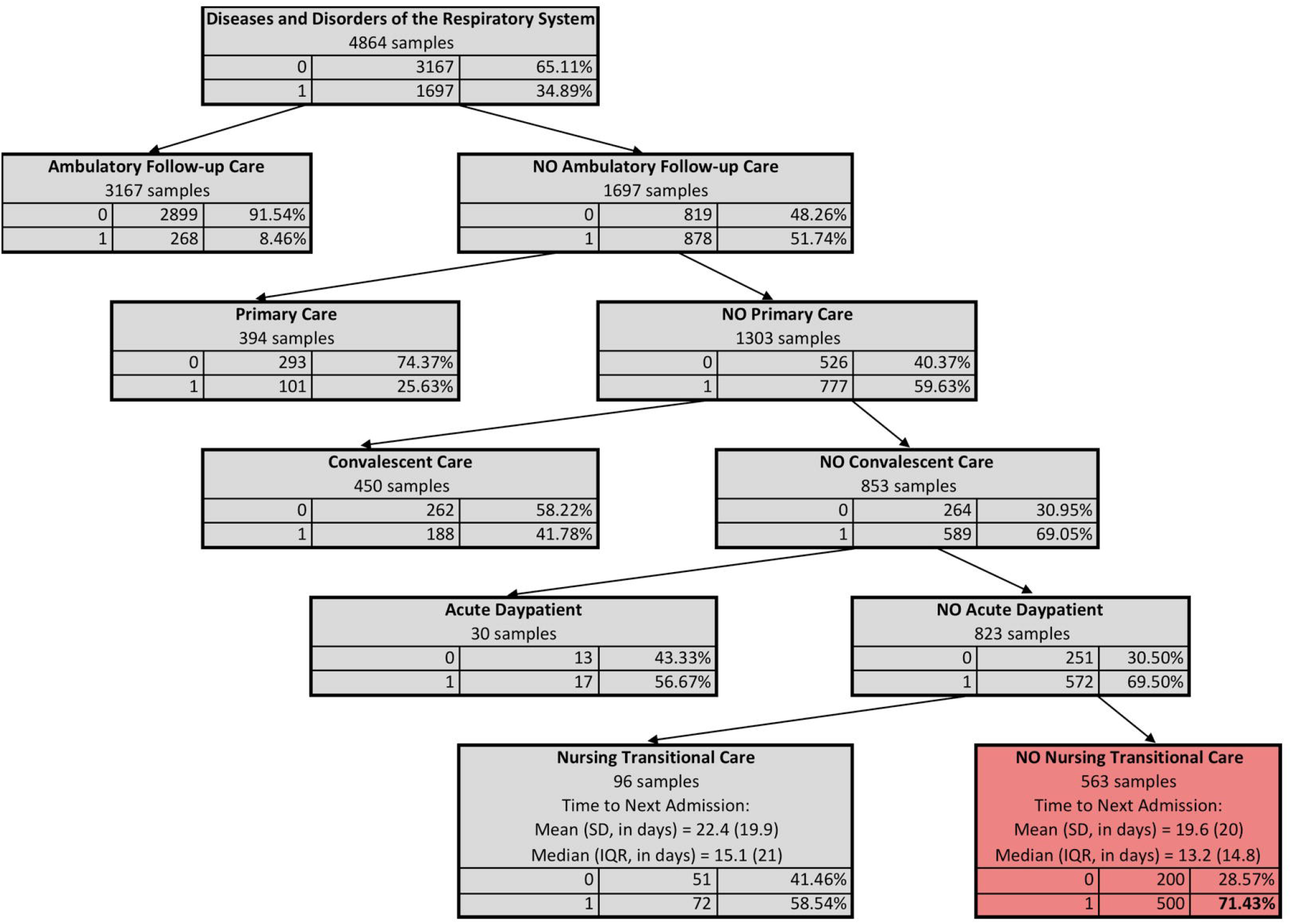

With schematic diagrams like Figures 2a and 2b, further analysis will be conducted to describe the findings for each segment of a study population. Although the pool of potential features includes many clinical and service-related (ambulatory, acute, post-acute, etc.) ones, the ML will only select important service features. For the illustrative analysis, in descending order of their marginal contributions to the 28-day re-hospitalization outcome, ambulatory follow-up care, primary care, and acute day patient care were selected to partition the respiratory segment of the 50-64 population. In contrast, ambulatory follow-up care, primary care, convalescent care, acute day patient care, and transitional nursing care were selected to partition the 65+ respiratory segment. After describing the relative orders with specifically selected services, the 28-day re-hospitalization outcome for a selected service will be analyzed and inferred. For instance, the following was consistently observed in Figures 2a and 2b, but it is not a mathematical necessity. The earlier a service is selected, the lower the corresponding service-benefited subgroup’s 28-day re-hospitalization rate is. The subgroup that lacked all selected services usually shows the highest rate.

Conducting the analysis with paired populations for comparison is recommended as well. The development of this protocol is aimed to recognize the group of patients that fall through the cracks of services and compare the same set of services’ impacts on different inpatient populations. For example, differences were also remarkable despite what is shared between the 50-64 and 65+ subgroups that lacked ML-selected services. Most of the 50-64 respiratory patients who lacked ML-selected services were diagnosed with malignant neoplasm of the respiratory system during both their index hospitalizations and 28-day re-hospitalizations. Meanwhile, most 65+ respiratory patients who lacked ML-selected services were diagnosed with chronic obstructive pulmonary diseases during their index hospitalizations and re-hospitalizations.

We will also benchmark the findings with Canadian Institute for Health Information (CIHI) standardized approaches. For example, the subgroups of 50-64 and 65+ respiratory patients who lacked ML-selected services also differ in CIHIs’ (2015) definition of resource-intensive multimorbidity and acute care interventions. The 50-64 respiratory patients who lacked ML-selected services were characterized by higher percentages of resource-intensive multimorbidity (20.71%) and acute care interventions (7.10%) than the following two groups. The first group was other 50-64 respiratory patients receiving ML-selected services (multimorbidity: 11.26%; resource-intensive intervention: 4.94%). The second group was the 65+ respiratory patients, regardless of whether ML-selected services were received (multimorbidity: received=14.78%, not received=14.03%; resource-intensive intervention: received=3.37%, not received=3.73%). Last but not least, we will further inspect the occurrences and timing of re-hospitalization for identified patient subgroups, besides 28-day re-hospitalization. For instance, compared to the two groups above, the 50-64 respiratory patients who lacked ML-selected services also showed a shorter re-hospitalization timeframe (median=12.80 days vs. 28.40 days for 50-64 who received the service, and 36.90 for all 65+).

Policy implications will be generated accordingly. While the wait time for post-acute care could be months, around 80% of those who received none of the ML-selected services were actually assigned the post-acute services, which could have prevented the re-hospitalization. Hence, our protocol will identify gaps in public medical systems and underscore the value of enabling what kind of access and its immediacy to post-acute care.

## ETHICS AND DISSEMINATION

### Research ethics approval

The research ethics for the illustrative example and ongoing territory-wide validation is approved and monitored by the HADCL and the Survey and Behaviour Research Ethics Committee of the Chinese University of Hong Kong. The forthcoming implementation and validation to segment inpatients for identifying tertiary prevention targets in the local healthcare system were approved by the regional ethical committee of HA. All patient information will be accessed, analyzed, and maintained in the HADCL and local medical offices without transferring to any places in order to protect confidentiality. All information was anonymized to avoid the re-identification problem. Access to data will only be granted to eligible researchers who sign and abide by the confidentiality agreements consistent with the ethics approvals.

### Dissemination

In addition to the concept-method alignment in studying the healthcare system with the ML pipeline we seek to disseminate via this protocol, the investigation results will be disseminated as scientific articles. Through presentations, meetings, and conferences, the investigators will communicate the results to the sponsors, i.e., HADCL and the local healthcare system, for further implementation. With re-hospitalization reduction being an objective of tertiary prevention, algorithms for profiling those with the highest re-hospitalization risk are planned to be integrated into hospitals’ CMS for screening to-be-discharge patients. Furthermore, taking a multiphasic screening approach, those screened at high risk of 28-day re-hospitalization will be targeted for in-depth assessments. The targeted in-depth assessment improves case identification and early intervention’s cost-effectiveness while enabling person-centered preventive measures with better integration with primary healthcare services.

The abovementioned disseminations will be summarized and reported back to the SPPR. Other relevant groups interested in the study results include social welfare practitioners or non-profit non-government agencies who run social services or participate in hospital transition programs designed for reducing hospitalization in the areas where we will conduct validations. The protocol advanced here is also implementable in different systems as part of the decision support mechanism to inform the venue-based sampling of patients with a high risk of re-hospitalization.

## DISCUSSION

The pressing need to gauge the effectiveness of ensembles of post-acute services has driven this ML-based analytic protocol. Including ML as a design element in protocols remains new in healthcare research. It is crucial to recognize the limitations of the ML-based approach and standardize the design as analytic protocols.

Compared with randomized control trials’ identification of homogeneous patients, the pool of potential candidates for the current protocol will be constructed by a clustering algorithm. The independence among features, including patient clinical profiles and services, will no longer be assumed in this protocol. Much of the clinical service assignment decisions are based on patients’ clinical profiles, with small but significant variations associated with the availability of services and individual differences among decision-makers. Consequently, the clinical homogeneity that enables the comparisons among different services’ effects on outcomes within each segment is only achieved in relative terms. Furthermore, the segmentation was based on a comprehensive array of clinical factors for parameterizing the differences in patients’ acute care utilization intensity to inform the practices of population case-mixing (CIHI, 2015). As a result, two patients sharing a segment may be similar across the multidimensional metrics of utilization intensity without sharing the same diagnosis, which is only one of the metrics.

The clinical homogeneity the current protocol seeks to establish with unsupervised ML may only reflect relative similarity in utilization intensity, parameterized in multidimensional clinical metrics. Homogeneous patient segments are intended to rid the confounds associated with the differences in patients’ intrinsic needs for the services by assessing services’ impact on patient outcomes. Nevertheless, the patients’ needs for services are also captured in our multidimensional measure of utilization intensity. Hence, assessing services’ effectiveness among patients with similar utilization intensity compared to the rest of the population is congruent with population-based planning that addresses the population’s service gaps.

The regression coefficients in linear models will not be explicitly found in the proposed protocol. The different services’ significantly different marginal impacts on 28-day re-hospitalization outcomes are only meaningful among clinically homogeneous patients. The ML-based effectiveness of selected services can only be interpreted in clinical profiling contexts. After all, what is considered critical lines of defense against 28-day re-hospitalization may differ for different clinical profiles, and so are their likelihood of having gaps.

## Supporting information

Checklist

## Data Availability

The datasets generated and/or analyzed during the current study are not publicly available due to restrictions being put on data sharing by Hong Kong Personal Data (Privacy) Ordinance (Cap. 486) (PDPO), including, but not exclusive to, PDPO Guidance Note in Cross-border Data Transfer. In addition, the Research Ethics Committees of the Hospital Authority do not allow a third-party transfer of patient data. Nor do the Ethics Committees permit study investigators to make public electronic health records of patients of Hospital Authority.

## FOOTNOTES

### Contributors

- The study concept and design were conceived by EL, AL & JG
- JG, CCC & OL conducted the data analysis.
- EL, AL & JG prepared the first draft of the manuscript,
- All were involved in article revisions for intellectual content.

